# The Omicron SARS-CoV-2 epidemic in England during February 2022

**DOI:** 10.1101/2022.03.10.22272177

**Authors:** Marc Chadeau-Hyam, David Tang, Oliver Eales, Barbara Bodinier, Haowei Wang, Jakob Jonnerby, Matthew Whitaker, Joshua Elliott, David Haw, Caroline E. Walters, Christina Atchison, Peter J. Diggle, Andrew J. Page, Deborah Ashby, Wendy Barclay, Graham Taylor, Graham Cooke, Helen Ward, Ara Darzi, Christl A. Donnelly, Paul Elliott

## Abstract

**Background:** The third wave of COVID-19 in England peaked in January 2022 resulting from the rapid transmission of the Omicron variant. However, rates of hospitalisations and deaths were substantially lower than in the first and second waves

**Methods:** In the REal-time Assessment of Community Transmission-1 (REACT-1) study we obtained data from a random sample of 94,950 participants with valid throat and nose swab results by RT-PCR during round 18 (8 February to 1 March 2022).

**Findings:** We estimated a weighted mean SARS-CoV-2 prevalence of 2.88% (95% credible interval [CrI] 2.76–3.00), with a within-round reproduction number (R) overall of 0.94 (0·91–0.96). While within-round weighted prevalence fell among children (aged 5 to 17 years) and adults aged 18 to 54 years, we observed a level or increasing weighted prevalence among those aged 55 years and older with an R of 1.04 (1.00–1.09). Among 1,195 positive samples with sublineages determined, only one (0.1% [0.0–0.5]) corresponded to AY.39 Delta sublineage and the remainder were Omicron: N=390, 32.7% (30.0–35.4) were BA.1; N=473, 39.6% (36.8–42.5) were BA.1.1; and N=331, 27.7% (25.2–30.4) were BA.2. We estimated an R additive advantage for BA.2 (vs BA.1 or BA.1.1) of 0.40 (0.36–0.43). The highest proportion of BA.2 among positives was found in London.

**Interpretation:** In February 2022, infection prevalence in England remained high with level or increasing rates of infection in older people and an uptick in hospitalisations. Ongoing surveillance of both survey and hospitalisations data is required.

**Funding:** Department of Health and Social Care, England.

Research in context

Evidence before this study
A search of PubMed using title or abstract terms (“Omicron” or “BA.1” or “BA.2”) and “prevalence” without language or other restrictions, identified 51 results (with no duplicates). All 51 results were evaluated, with 18 deemed relevant. One study focused on Omicron case rates in South Africa during the early stage after the discovery of the new variant (November 2021), one described genomic surveillance of SARS-CoV-2 in the USA (June – December 2021), one analysed clinical outcomes based on health records (January – December 2021), one described the results of whole-genome sequencing of SARS-CoV-2 samples collected in North Africa (March – December 2021), and one was from a previous REACT survey round (November – December 2021). The others focused on the mutation distribution of Omicron, disease severity, immune response, vaccine effectiveness, and prevalence in animal hosts.
Added value of this study
We analysed data from throat and nose swabs collected at home by a randomly selected sample of residents of England, aged 5 years and older, obtained during round 18 (8 February to 1 March 2022) of the REal-time Assessment of Community Transmission-1 (REACT-1) study. We estimated a weighted prevalence of SARS-CoV-2 of 2.88% (95% CrI 2.76–3.00) in England in February 2022, which was substantially lower than that estimated in January 2022 (4.41% [4.25–4.56]). The within-round dynamics differed by age group with weighted prevalence falling among children (aged 5 to 17 years) with an R of 0.79 (0.74–0.84) and adults aged 18 to 54 years with an R of 0.92 (0.89–0.96), in contrast to the level or increasing weighted prevalence among those aged 55 years and older with an R of 1.04 (1.00–1.09). Exponential models estimated a daily growth rate advantage of 0.12 (0.11–0.13) in the odds of BA.2 (vs BA.1 or BA.1.1) corresponding to an R additive advantage of 0.40 (0.36–0.43).
Implications of all the available evidence
Random community surveys of SARS-CoV-2 provide robust insights into transmission dynamics and identify groups at heightened risk of infection based on estimates of population prevalence that are unbiased by test-seeking behaviour or availability of tests. In England, replacement by BA.2 of other Omicron sublineages, the level or increasing rates of infection in older people and the uptick in hospitalisations in England toward the end of February 2022 require ongoing surveillance, both to monitor the levels of current (and future) SARS-CoV-2 variants and the risks of severe disease.

## Introduction

As part of the government’s plan for “living with COVID-19”^1^, domestic legal restrictions concerning SARS-CoV-2 infection ended in England on 24 February 2022^2^, fourteen months after the UK began its national SARS-CoV-2 vaccination programme^3^. The removal of legal restrictions came just over a month after the third-wave peak in deaths within 28 days of a positive SARS-CoV-2 test in England, and the rapid rise in transmission related to the Omicron variant.^4^ These occurred despite high levels of vaccine coverage in adults and children aged 12 years and older^5^ but with substantially lower peak rates of hospitalisations and deaths than in the first and second waves in April 2020 and January 2021, respectively^6^.

Changes in population-wide testing availability and policies have complicated interpretation of the routine SARS-CoV-2 testing data. These include the fact that those testing positive using a lateral flow device (LFD) are no longer required to take confirmatory PCR tests^7^, variable test-seeking behaviour and periods when the demand for testing (LFD and PCR) exceeded supply. Reported cases now include those that were LFD-positive only, PCR-positive only and positive on both types of test, although it is unclear what proportion of people who undertake a home-test LFD report the results, and most asymptomatic infections go unreported. In addition, from 31 January 2022, the national reporting includes reinfections for the first time^8^.

Against this backdrop, random community surveys of SARS-CoV-2 provide situational awareness based on unbiased estimates of population prevalence^9^. The REal-time Assessment of Community Transmission-1 (REACT-1) study has obtained throat and nose swabs from random samples of the population of England (5 years of age and older) approximately monthly since May 2020, giving estimates of the community prevalence of swab-positivity in England and insights into the underlying transmission dynamics. Here we report the results from the eighteenth round of data collection of the REACT-1 study, in which samples were collected from 8 February to 1 March 2022.

## Methods

The REACT-1 study has conducted cross-sectional surveys of random samples of the population of England at ages 5 years and older^10^ over two-to three-week periods on a monthly basis since May 2020 (exceptions were December 2020 and August 2021). Participants have provided a self-administered throat and nose swab which was then analysed for SARS-CoV-2 by reverse transcription polymerase chain reaction (RT-PCR). A positive test result was recorded if both N gene and E gene targets were detected or if N gene was detected with cycle threshold (Ct) value below 37. We focus here on results from round 18 (8 February to 1 March 2022) involving N=94,950 participants with a valid RT-PCR test result (including 685 samples [18 positives] obtained between 2 and 4 March 2022), and compare them to results from round 17 (5 to 20 January 2022, including 862 samples [36 positives] obtained between 21 and 24 January 2022). Our sampling frame was based on the National Health Service (NHS) general practitioner list of patients in England. Participants completed a brief registration and an online or telephone questionnaire from which we collected information on age, sex, residential postcode, ethnicity, household size, occupation, potential contact with a COVID-19 case, symptoms and other variables^11^. We used the postcode of residence to link to an area-level Index of Multiple Deprivation^12^ and urban/rural status.^13^

As previously described^14,15^, to the beginning of May 2021, we obtained random samples with approximately equal numbers of people in each lower-tier local authority area (LTLA) in England, but from round 12 (20 May to 7 June 2021), to improve sample representativeness, we changed our sampling procedure to be in proportion to population size at LTLA level. Following a successful pilot study in round 14 (9 to 27 September 2021), from round 15 (19 October to 5 November) onward we switched from collecting dry swabs by courier to ‘wet’ (saline) swabs sent to the laboratory by priority post. We included a small proportion of samples obtained after the nominated closing date for that and each subsequent round to account for postal delays in the return of swabs. From round 16, (23 November to 14 December 2021) we included a multiplex for influenza A and B in addition to SARS-CoV-2 (only the results for SARS-CoV-2 are presented here). In round 18, we added an incentive to increase response rates among under-represented groups. For returning their completed test, those aged 13 to 17 and 35 to 44 were offered a gift voucher worth £10 while those aged 18 to 34 were offered a gift voucher worth £20. We used random iterative method (rim) weighting^16^ to provide prevalence estimates for the population of England as a whole, adjusting for age, sex, deciles of the Index of Multiple Deprivation, LTLA counts, and ethnic group.

Samples testing positive with Ct values of 34 or less in the N gene and/or the E gene were sent for viral genome sequencing to the Quadram Institute, Norwich, UK. We used the ARTIC protocol^17^ (version 4.1) for viral RNA amplification, CoronaHiT for preparation of sequencing libraries,^18^ the ARTIC bioinformatics pipeline^17^ and assigned sublineages using PangoLEARN (version 2022-02-02).^19^ Sequencing data for the present analyses were available for positive swabs obtained up to 21 February 2022.

### Data analyses

For each round, we estimated weighted prevalence and 95% credible intervals overall and by socio-demographic and other variables. We used logistic regression to estimate the odds of testing positive by employment, ethnicity, household size, children in household, urban status, and deprivation, adjusting for age, region and the other variables examined.

We fit a Bayesian logistic regression model to the proportion of BA.2 sublineage compared to BA.1 or BA.1.1 during round 18 to investigate whether there was a daily growth rate advantage for the odds of BA.2 versus BA.1 or BA1.1. The daily percentage growth in the odds of BA.2 infection was estimated from the exponential of the daily growth rate. The estimated additive R advantage was estimated as the daily growth rate advantage multiplied by the Omicron-specific mean generation time of 3.3 days.^20^

Using a non-parametric Kruskal-Wallis test, we compared the distribution of the Ct values among test-positive swabs (N gene and E gene where Ct>0) across sublineages and, within each determined sublineage, by symptoms status: in those declaring any symptoms or any of the four ‘classic’ COVID-19 symptoms (defined as loss or change of sense of smell or taste, fever, new persistent cough) vs those not declaring any symptom in the month prior to swabbing.

To visualise temporal trends in SARS-CoV-2 swab positivity, we applied a Bayesian penalised-spline (P-spline) model^21^ to daily weighted prevalence data. Specifically, we used a No-U-Turn Sampler in logit space and data were split into approximately 5-day sections by regularly spaced knots. Edge effects were minimised by adding further knots beyond the study period. We used fourth-order basis splines (b-splines) over the knots including a second-order random-walk prior distribution on the coefficients of the b-splines to guard against overfitting; the use of this prior distribution penalised changes in the growth rate unless supported by the data.^15,22^ We also fit P-splines separately to three broad age groups (17 years and under, 18 to 54 years, 55 years and older) with the smoothing parameter obtained from the model fit to the entire dataset.

To investigate growth or decay of daily weighted prevalence between round 17 and round 18, we used an exponential model assuming a binomial distribution for the number of positives out of the total number of samples per day. To estimate the growth rate and posterior credible intervals, we used day of sampling where reported (otherwise day of first scan of the swab by the Post Office if available) with a bivariate No-U-Turn Sampler and uniform prior distribution for the probability of swab positivity.^23^ We estimated the reproduction number R from the daily growth rate *r* from

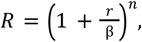

where *r* is the exponential growth (or decay) rate, and the shape parameter *n* and the rate parameter *β* were set to 0.89 and 0.27, respectively ensuring under a gamma distribution assumption, a mean generation time of 3.3 days and a standard deviation of 3.5 days, as previously reported.^20^

We estimated a neighbourhood smoothed prevalence at the LTLA level by randomly selecting within each LTLA 15 participants and calculating the prevalence of infection among the nearest M people, where M was the median number of study participants within 30 km of these participants.

We used R software^24^ for the data analyses.

### Ethics

We obtained research ethics approval from the South Central-Berkshire B Research Ethics Committee (IRAS ID: 283787).

## Results

### SARS-CoV-2 swab-positivity prevalence in February 2022

A total of 635,000 individuals were invited to participate in round 18 of the REACT-1 study; 133,118 (20.96%) registered and were sent a RT-PCR kit and 94,950 (15.0%) returned the swab and had a valid RT-PCR test result. Swabs were obtained from 8 February to 1 March 2022 (including 685 swabs and 18 positives obtained between 2 to 4 March 2022) (Figure 1); 2,731 tested positive for SARS-CoV-2, yielding a weighted prevalence of 2.88% (95% CrI 2.76–3.00) (Table S1). This was the second highest weighted prevalence observed throughout the REACT-1 study after that in round 17 (5 to 20 January 2022; N=862 further samples (including 36 positives) from 21-24 January 2022) which was 4.41% (4.25–4.56), and was almost twice the weighted prevalence observed in January 2021 (round 8) at 1.57% (1.49–1.66). During round 18 incentives were used, which increased the response rates overall from 12.2% in round 17 to 15.0% in round 18 despite lower swab return rates in round 18 (70.8%) compared to round 17 (73.0%). Response rates more than doubled in those aged 13 to 17 years and more than tripled in those aged 25 to 34 years, which were previously under-represented.

**Figure 1.**
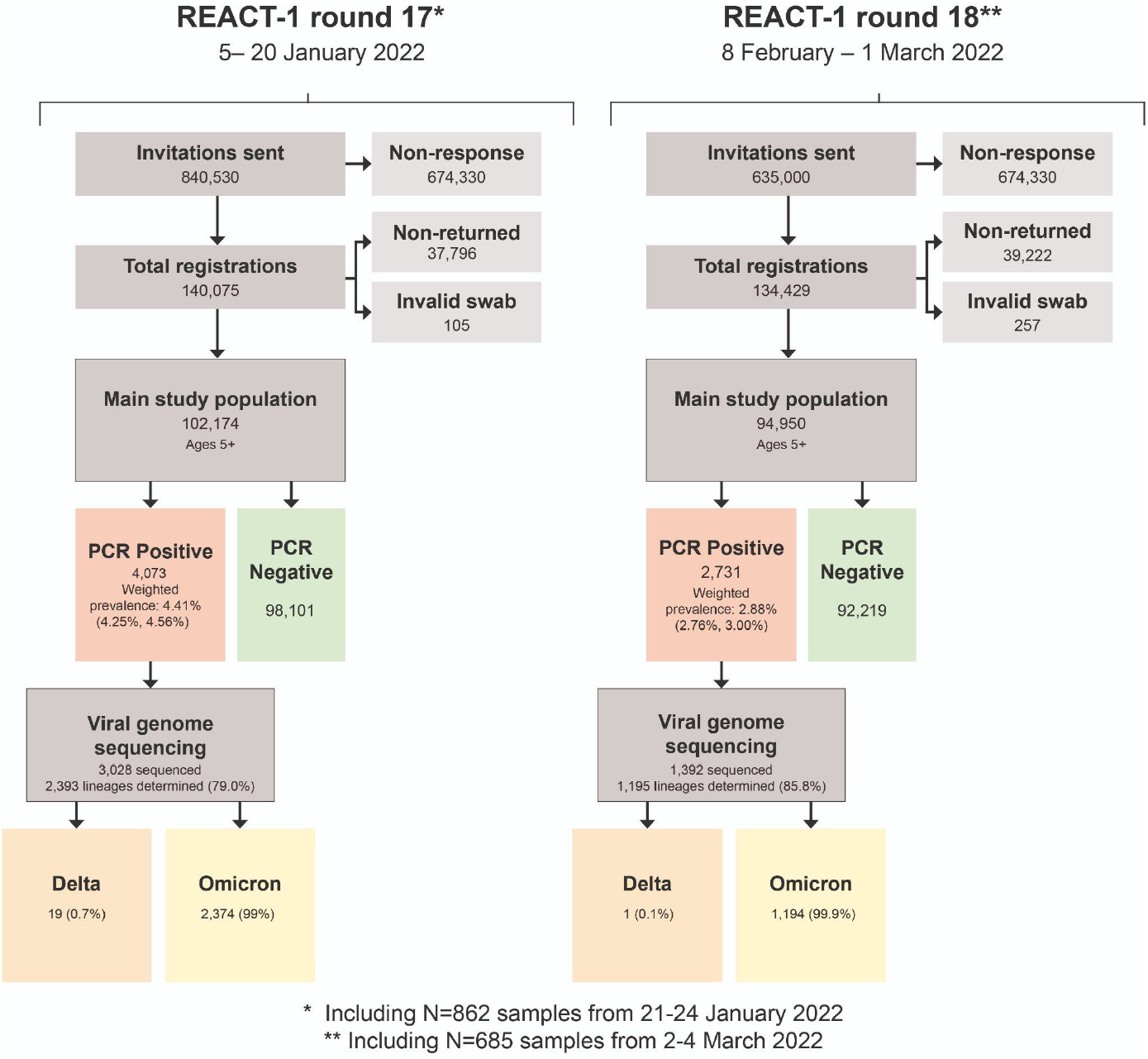
Flow chart showing numbers of participants in round 17 (5-20 January 2022) and round 18 (8 February - 1 March 2022) of REACT-1.

### Omicron sublineages

We determined a total of 1,195 sublineages (85.6%; 95% CI 83.9–87.6) from the 1,392 positive samples obtained to 21 February 2022 with sequencing data (Table 1). Of these one (0.1%; 95% CI 0.0–0.5) corresponded to AY.39 Delta sublineage and was detected in a participant living in East of England who did not report having travelled within or outside the UK in the two weeks prior to swabbing. All other sublineages were Omicron variant and included 32.7% (95% CI 30.0–35.4; N=390) BA.1; 39.6% (36.8–42.5; N=473) BA.1.1 and 27.7% (25.2–30.4; N=331) BA.2 sublineages.

**Table 1.**
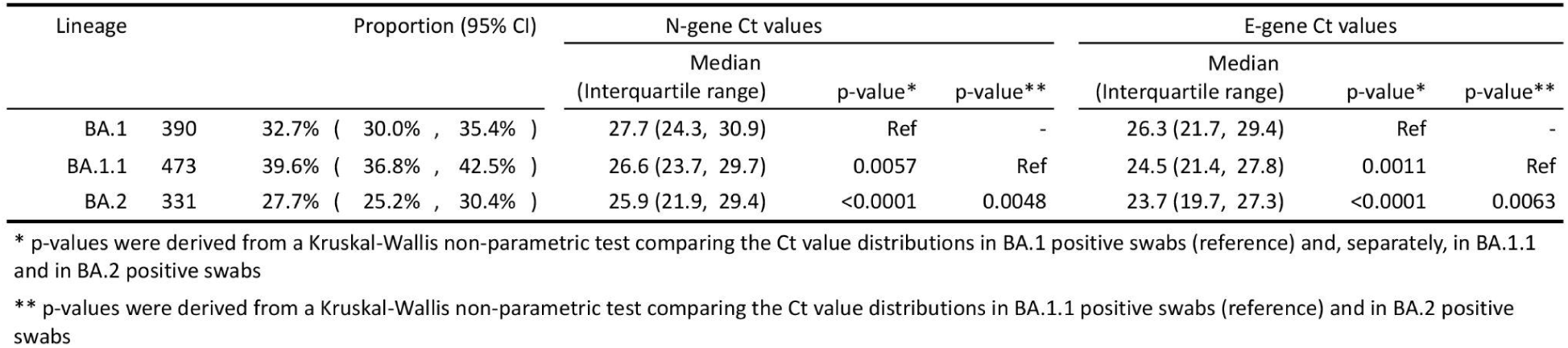
Proportion of each of the N=1,194 SARS-CoV-2 Omicron sublineages detected in positive samples obtained to 21 February 2022 with at least 50% genome coverage from round 18. One Delta sublineage (AY.39) was also determined. Results are based on 1,392 positive sequenced samples. For each Omicron sublineage, distribution of Ct values is summarised by its median and interquartile range, and distributions are compared across sublineages using a non-parametric Kruskal-Wallis test. Corresponding p-values are reported.

We estimated a daily growth rate advantage of 0.12 (95% CrI 0.11, 0.13) in the odds of BA.2 (vs BA.1 or BA.1.1) and a proportion of BA.2 of 47.2% (42.8–51.7) on 21 February 2022 (Figure 2A). Assuming a mean generation time of 3.3 days, this corresponds to an R additive advantage for BA.2 (vs BA.1 or BA.1.1) of 0.40 (95% CrI 0.36–0.43). Characteristics of swab positive participants with BA.1, BA.1.1 and BA.2 sublineages were similar except for an excess of BA.2 infections in London compared to other regions. Specifically, of the 212 positive swabs with determined sublineages from participants living in London to 21 February 2022, all were Omicron and 94 (44.3%; 95% CI 37.8–51.1) were BA.2 (Figure 2B).

**Figure 2-A.**
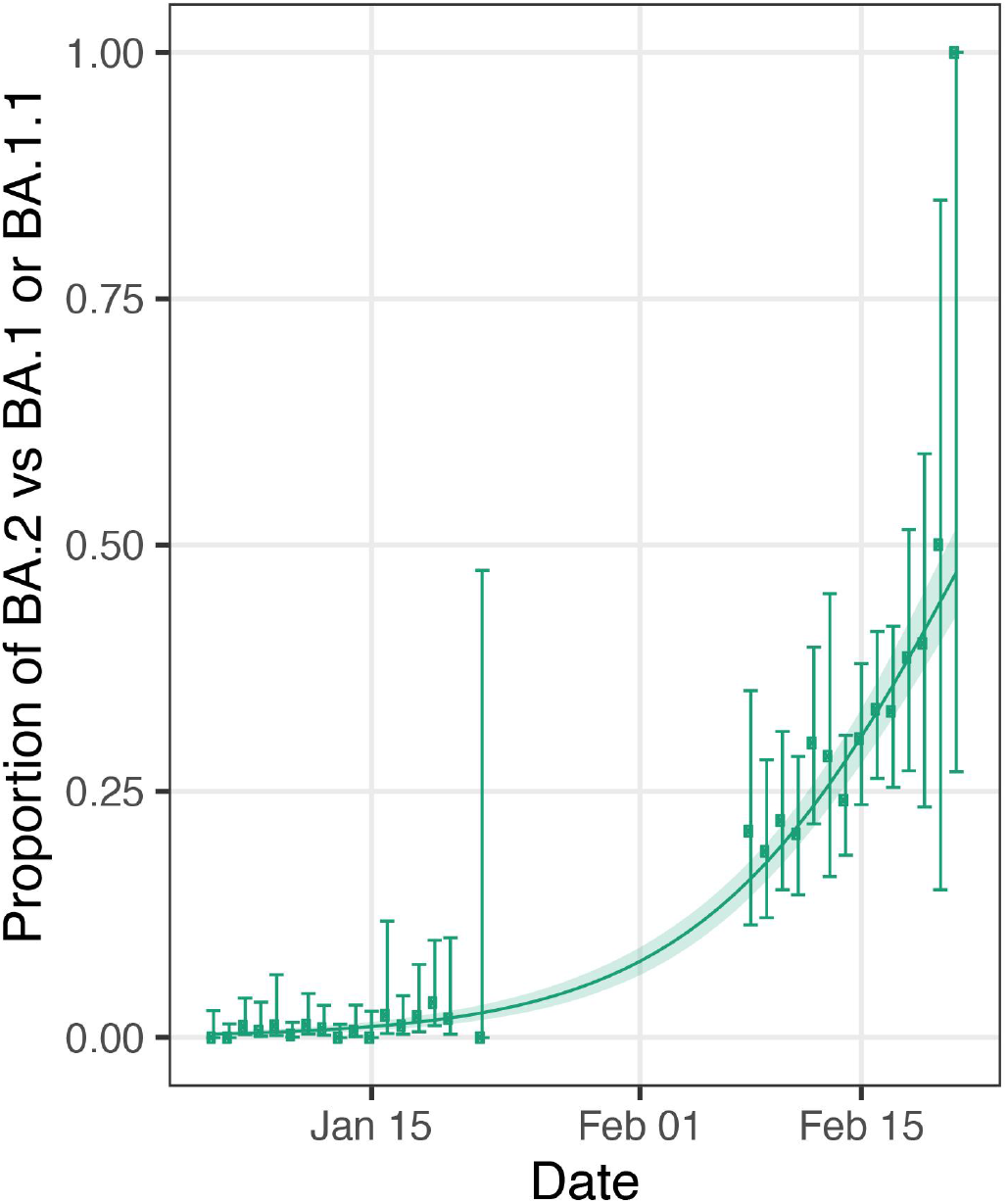
Daily proportion of BA.2 (vs BA.1 or BA.1.1) infections among positive swabs with determined lineage and at least 50% genome coverage in round 17 and round 18 for samples obtained to 21 February 2022. Point estimates are represented (dots) along with 95% confidence intervals (vertical lines). Smoothed estimates of the proportion are also shown (solid line) together with their 95% credible intervals (shaded regions)

**Figure 2B.**
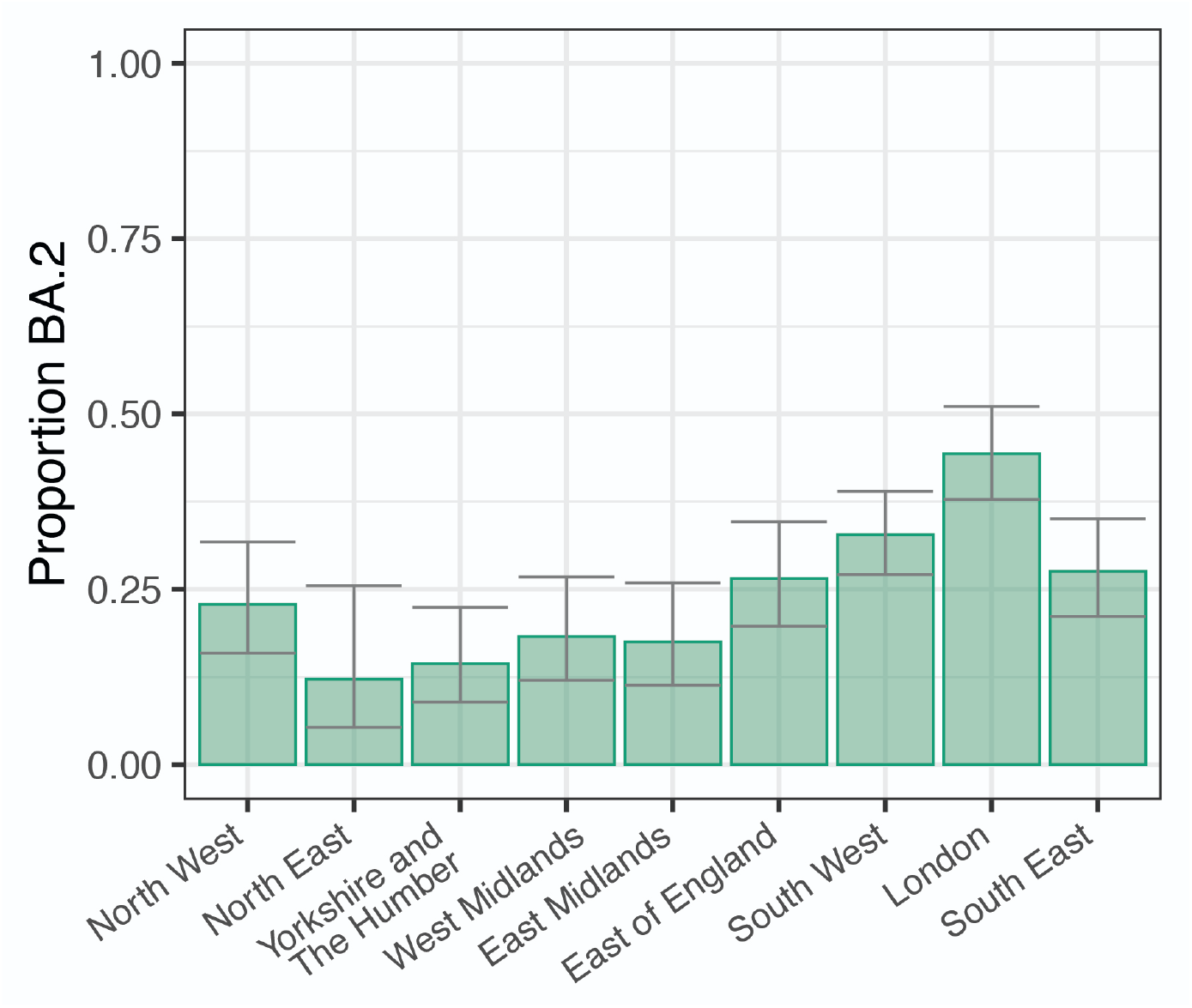
Regional proportion of BA.2 infections among positive swabs with determined lineage and at least 50% genome coverage in round 18 for samples obtained to 21 February 2022. Point estimates are represented (bars) along with 95% confidence intervals (vertical lines)

**Figure 2C.**
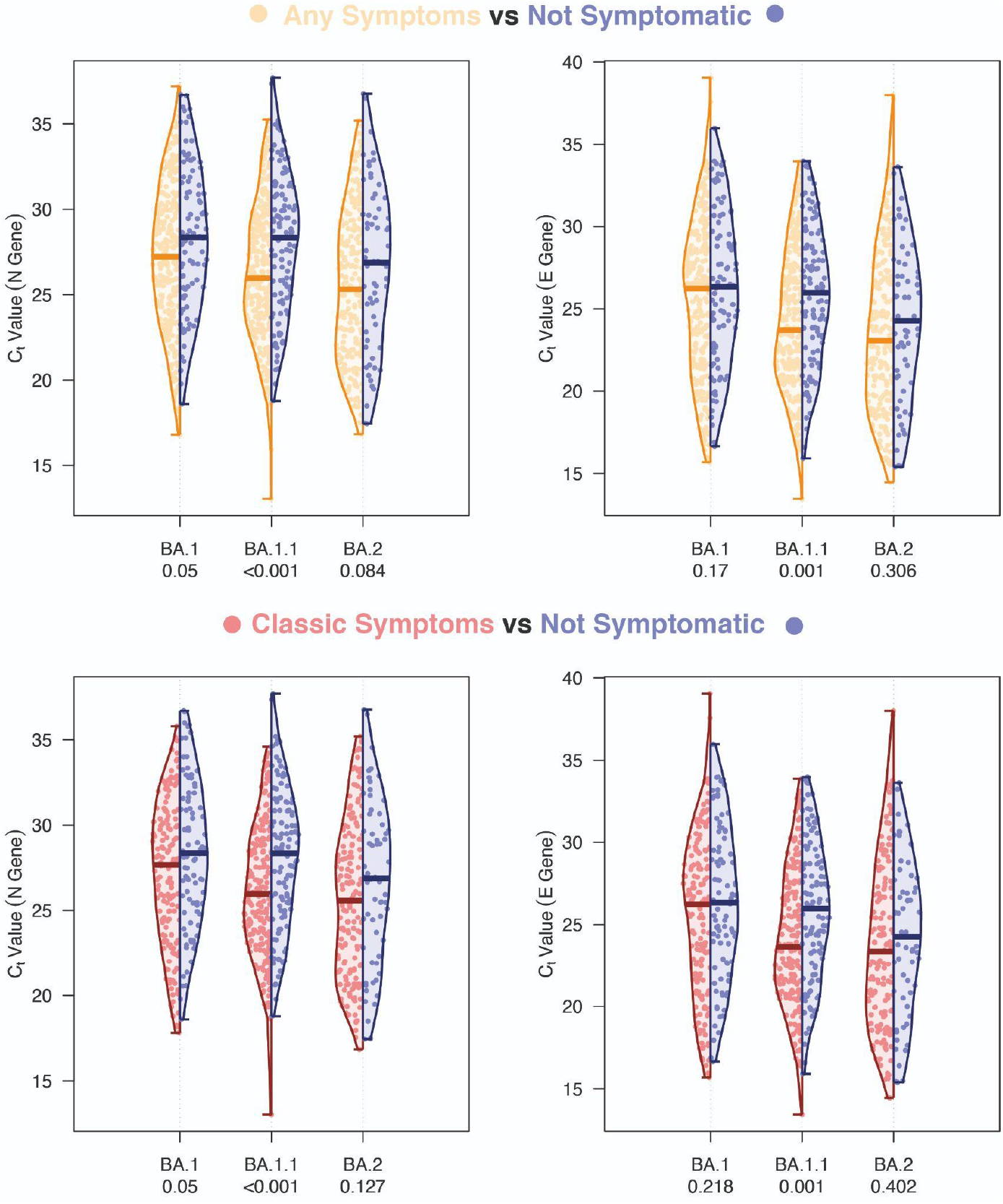
Distribution of Ct values for the N gene (left) and E gene (right) in swab-positive samples from round 18 obtained to 21 February 2022. For each of the three determined Omicron sublineages (BA.1, BA.1.1 and BA2) separately, distributions of Ct values are plotted for those reporting at least one symptom in the month prior to swabbing (orange), in those reporting any of the classic symptoms – loss or change of sense of smell or taste, fever, new persistent cough (red) – and those not reporting any symptoms (blue). Comparison of the distribution of Ct values in those reporting symptoms (any or at least one of the classic four COVID-19 symptoms) and those not reporting any symptoms was done using a Kruskal-Wallis test, and we report the corresponding p-value.

N gene Ct values for positive swabs with BA.2 sublineage (median 25.9, interquartile range 21.9–29.4) were lower (Kruskal Wallis test p-value <0.005) than in positive swabs with BA.1 (median 27.7, interquartile range 24.3–30.9) and BA.1.1 (median 26.6, interquartile range 23.7–29.7) (Table 1). N gene Ct values in positive swabs with BA.1.1 were lower than in positive swabs with BA.1 (p=0.006). Similar results were observed using E gene Ct values with median Ct values of 26.3 (interquartile range 21.7–29.4), 24.5 (interquartile range 21.4, 27.8), and 23.7 (interquartile range 19.7–27.3) in positive swabs with BA.1, BA.1.1, and BA.2, respectively.

For both N gene and E gene, we found lower Ct values in swab positive samples with BA.1.1 in those reporting any or ‘classic’ (loss or change of sense of smell or taste, fever, new persistent cough) symptoms in the month prior to swabbing compared to those not reporting symptoms (p <0.001) (Figure 2C). Differences in Ct values by symptom reporting for positive swabs with BA.2 were less certain.

### Dynamics of SARS-CoV-2 swab-positivity

P-spline models estimated for the whole of REACT-1 showed a decreasing prevalence between round 17 (5 to 20 January 2022) and round 18 (8 February to 1 March 2022) overall and within round 18 (Figure 3A); an exponential model estimated an overall between-round R (round 17 to round 18) of 0.96 (0.96–0.96) and a within-round 18 R of 0.94 (0.91–0.96) both with lower than 0.01 posterior probability that R>1 (Table S2).

**Figure 3A.**
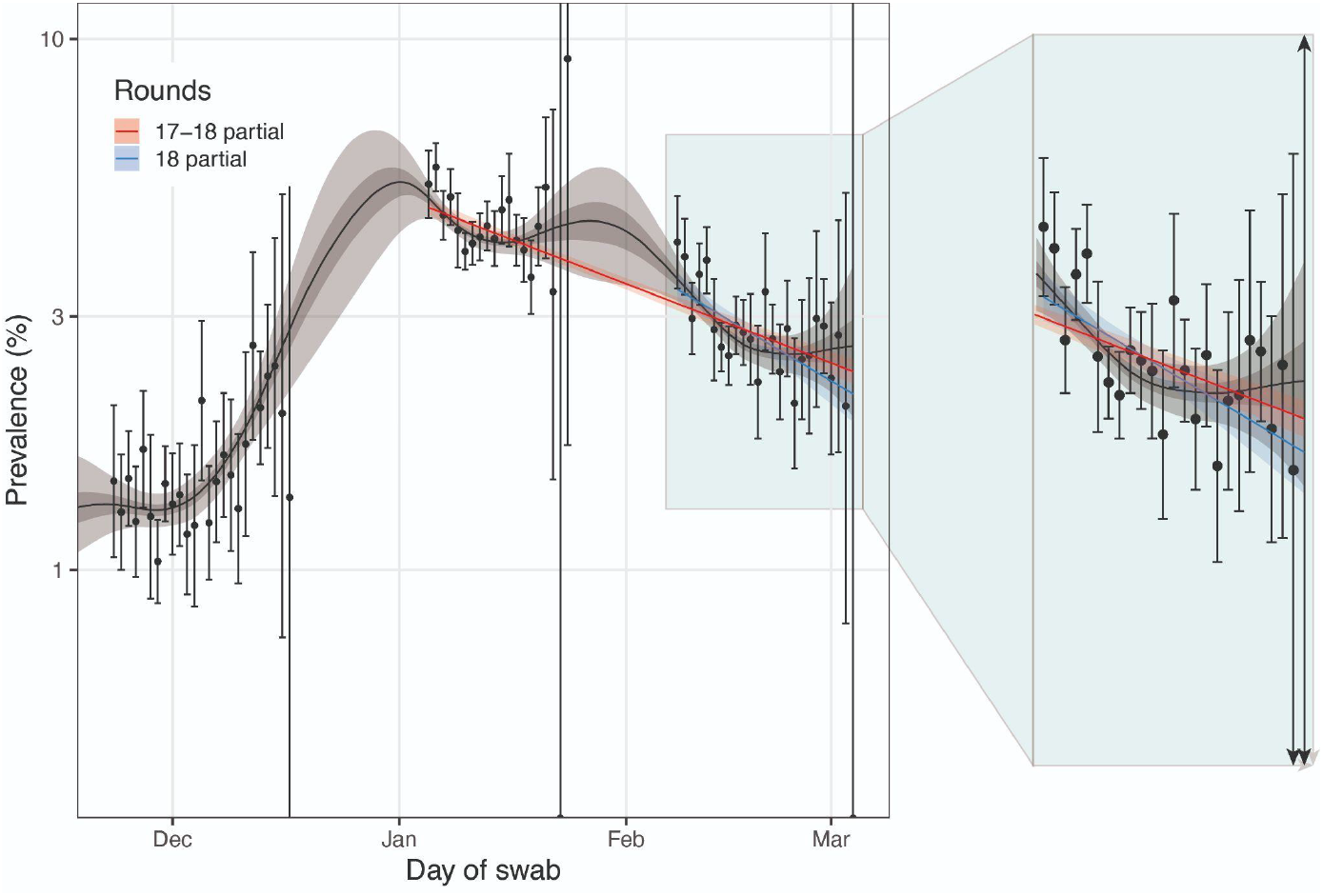
Comparison of an exponential model fit to round 18 (blue), an exponential model fit to round 17 and round 18 (red) and a P-spline model fit to all rounds of REACT-1 (black, shown here only for rounds 16, 17 and 18). Shaded red region shows the 95% posterior credible interval for the exponential models, and the shaded grey region shows 50% (dark grey) and 95% (light grey) posterior credible interval for the P-spline model. Results are presented for each day (X axis) of sampling for round 16, round 17 and round 18 and the weighted prevalence of swab-positivity is shown (Y axis) on a log scale. Weighted observations (black dots) and 95% confidence intervals (vertical lines) are also shown.

**Figure 3B.**
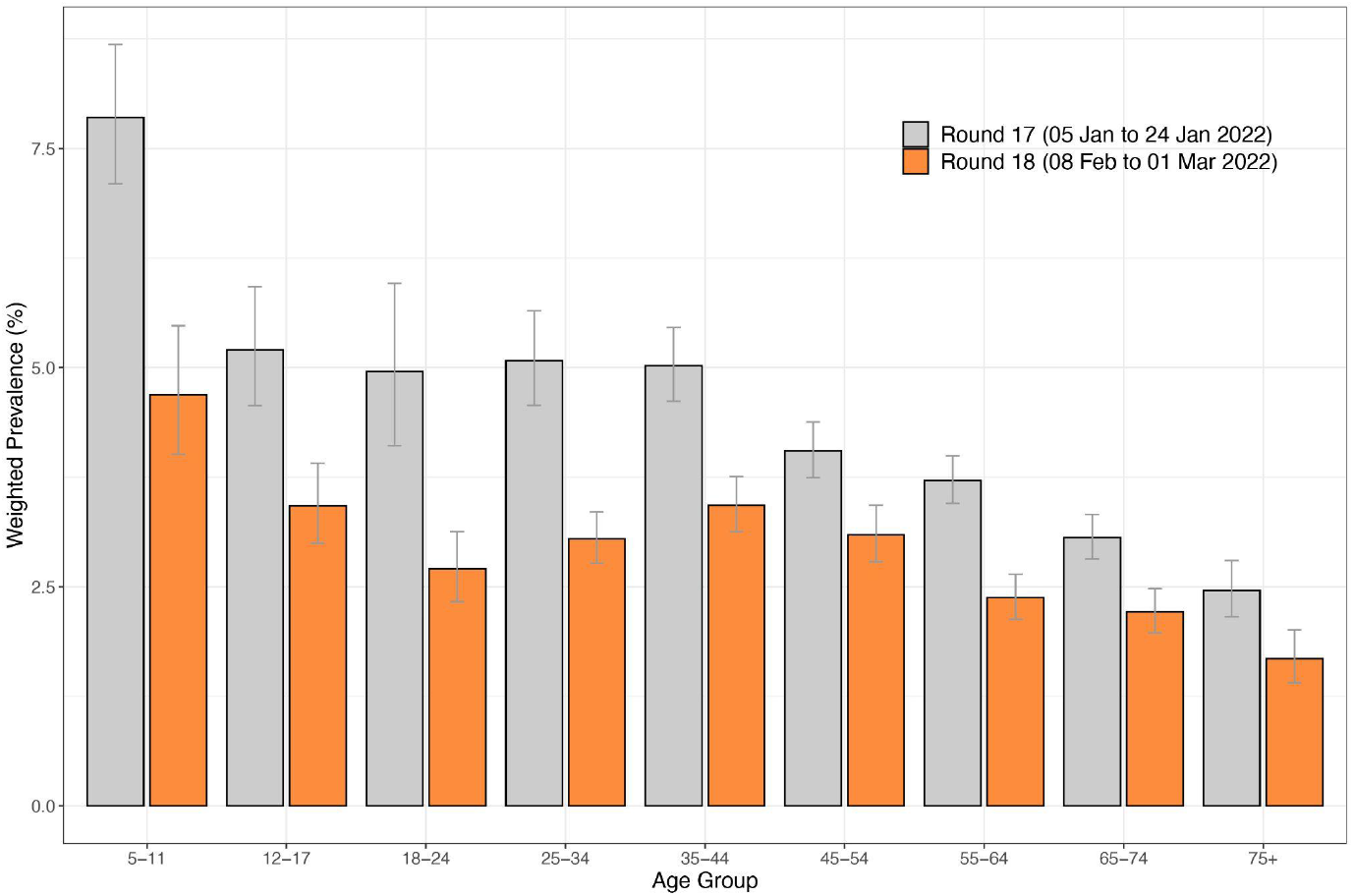
Weighted prevalence of swab-positivity by age group for round 17 and round 18. Bars show the prevalence point estimates (grey for round 17 and orange for round 18), and the vertical lines represent the 95% confidence intervals.

**Figure 3C.**
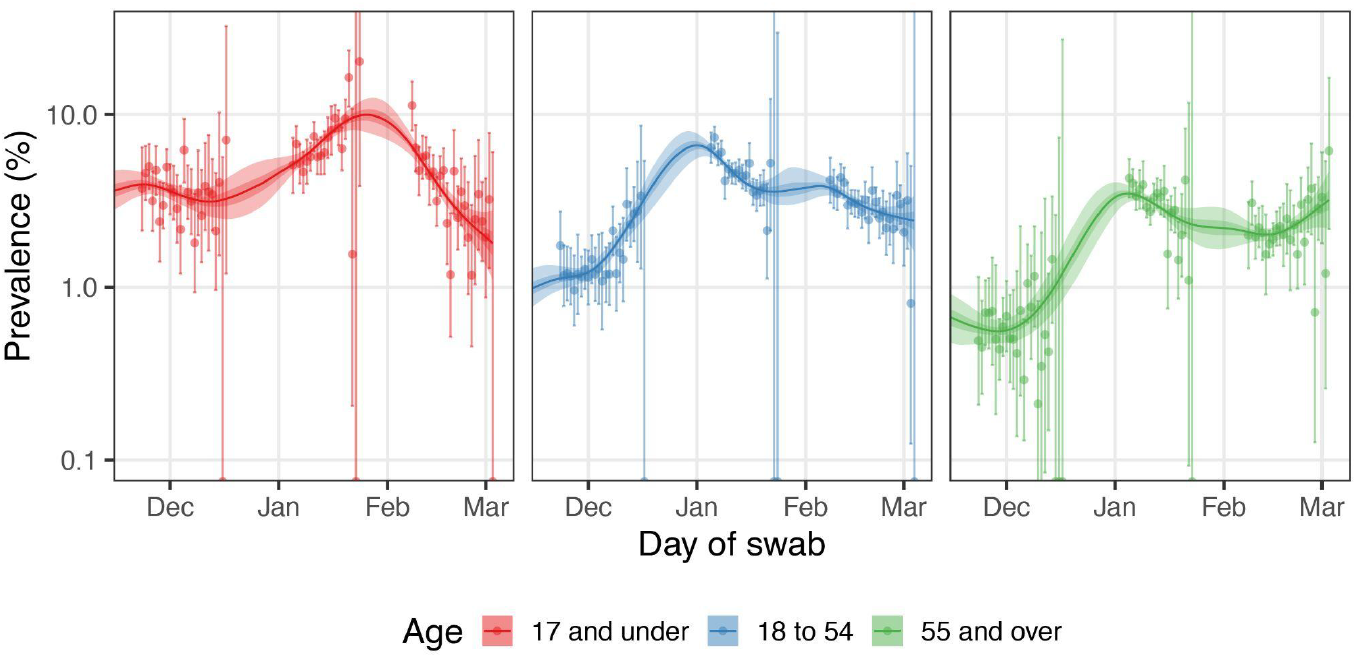
Comparison of P-spline models fit to SARS-CoV-2 swab-positivity data from all rounds of REACT-1 for those aged 17 years and under (red), those aged 18 to 54 years inclusive (blue) and those aged 55 years and over (green). Shaded regions show 50% (dark shade) and 95% (light shade) posterior credible interval for the P-spline models. Results are presented for each day (X axis) of sampling for round 16, round 17, and round 18 and the prevalence of swab-positivity is shown (Y axis) on a log scale. Weighted observations (dots) and 95% confidence intervals (vertical lines) are also shown.

**Figure 3D.**
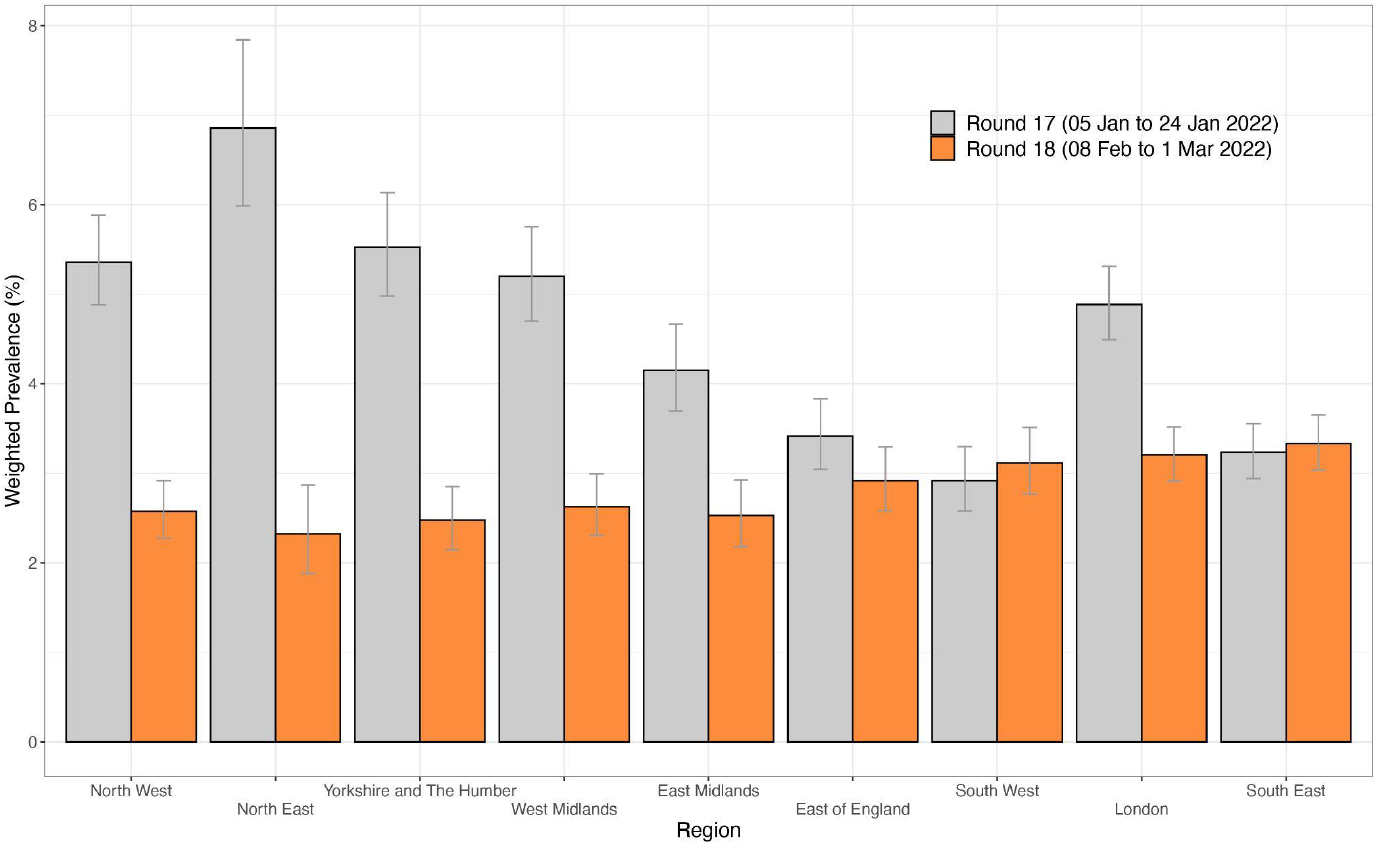
Weighted prevalence of swab-positivity by region for round 17 and round 18. Bars show the prevalence point estimates (grey for round 17 and orange for round 18), and the vertical lines represent the 95% confidence intervals.

**Figure 3E.**
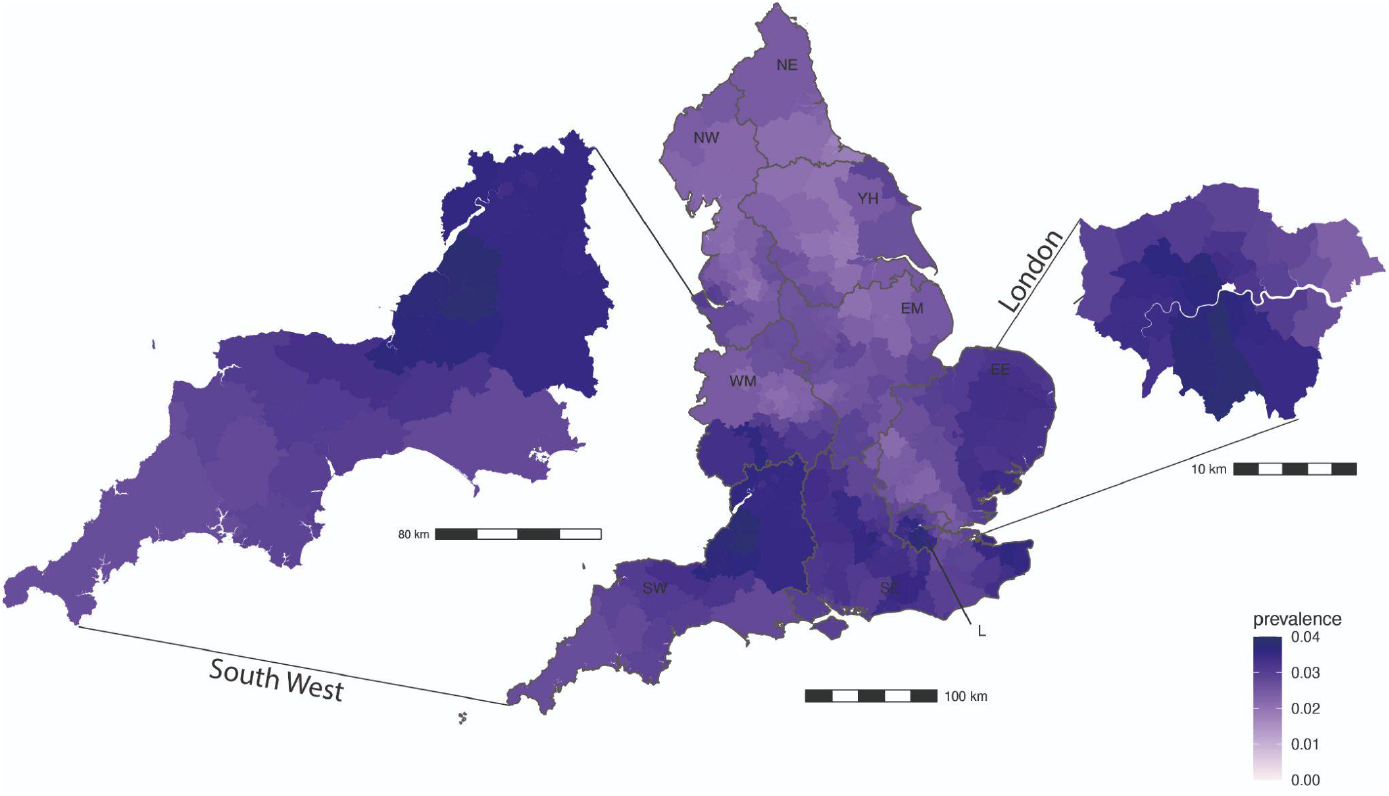
Neighbourhood smoothed average SARS-CoV-2 swab-positivity prevalence by lower-tier local authority area for round 18. Neighbourhood prevalence calculated from nearest neighbours (the median number of neighbours within 30 km in the study). Average neighbourhood prevalence displayed for individual lower-tier local authorities for the whole of England. Regions: NE = North East, NW = North West, YH = Yorkshire and The Humber, EM = East Midlands, WM = West Midlands, EE = East of England, L = London, SE = South East, SW = South West

Weighted prevalence in round 18 by age ranged from 1.68% (1.41–2.01) in those aged 75 years and older to 4.69% (4.01–5.48) in those aged 5 to 11 years. Weighted prevalence in round 18 was higher in the youngest age group compared to all other ages (Table S3A, Figure 3B). At all ages, weighted prevalence in round 18 was lower than that observed in round 17 (Figure 3B) with estimated between-round R of 0.96 (0.95–0.97) in those aged 17 years and under, 0.96 (0.95–0.96) in those aged 18 to 54 years, and 0.96 (0.96–0.97) in those aged 55 years and older, all with <0.01 posterior probability that R>1 (Table S2). Weighted prevalence also fell during round 18 among those aged 5 to 17 years with within-round R of 0.79 (0.74–0.84) and those aged 18 to 54 years with within-round R of 0.92 (0.89–0.96) and <0.01 posterior probability that R>1. However, within-round 18 trends in those aged 55 years and older suggested a level or increasing prevalence with within-round R of 1.04 (1.00–1.09) and 0.96 posterior probability that R>1 (Figure 3C).

Weighted prevalence fell in all regions between round 17 and round 18 (with <0.01 posterior probability that between-round R>1) except in South East and South West (Tables S2-S3A, Figure 3D). Weighted prevalence in round 18 remained high, ranging from 2.33% (1.88–2.87) in North East to 3.20% (2.92–3.52) in London and 3.33% (3.04–3.66) in South East (Table S3A, Figure 3D). At lower-tier local authority level, of the 10 highest smoothed estimates of prevalence based on a nearest neighbour method, six were in London (Lambeth, Croydon, Sutton, Merton, Camden, Wandsworth) and four in South West (Bath and North East Somerset, City of Bristol, South Gloucestershire, North Somerset), (Figure 3E).

### Determinants of SARS-CoV-2 swab-positivity

We found higher weighted prevalence in (i) larger households compared to single-person households, (ii) households with one or more children compared to households without children, (iii) those having been in contact with a (confirmed or suspected) COVID-19 case compared to those without such contact, (iv) those not shielding, and (v) those reporting ‘classic’ COVID-19 or any symptoms in the month prior to swabbing compared to those not reporting any symptoms (Table S3B). In multivariable logistic regression we found elevated risk of swab positivity in healthcare or care home workers (vs other non-essential workers) with mutually adjusted OR of 1.18 (1.02–1.36), and in participants living in households with one or more children (vs household without children) with mutually adjusted OR of 1.35 (1.19–1.53) (Table S4).

## Discussion

This eighteenth round of the REACT-1 study during February 2022 was undertaken following the rapid and almost complete replacement of Delta by Omicron variant in England in December 2021 and January 2022. Infection rates remain higher than recorded in REACT-1 at any time during the pandemic except at the peak of Omicron infections in early January 2022, and are almost double those observed in January 2021 during the second wave.

Although infection rates fell overall in February 2022, there was a concerning trend of level, or possibly increasing, prevalence at older ages (55 years and older) during round 18. This may feed through into higher rates of symptomatic infections and hospitalisations among this vulnerable group, despite the protection conferred by their high levels of vaccination. Notably, following a decline in hospitalisations in England from a peak around 9 January 2022, hospitalisations have begun to rise again from around 26 February, toward the end of round 18 ^25^. It will be important to continue to monitor infections, and hospitalisations, among older people, many of whom will have received a booster dose of vaccine 4 to 5 months earlier, with evidence suggesting waning of immunity a few weeks post booster dose ^26^. In this regard, the UK government has announced a fourth dose of vaccine will be offered to vulnerable people, including those over the age of 75 years, from spring 2022^27^, following a similar policy of administration of fourth doses in Israel^28^. In addition, the government’s removal of all domestic legal restrictions concerning COVID-19 in England on 24 February 2022^2^ was widely anticipated in the weeks before, which may have led to increased population mixing across ages, and contributed to the observed infection and hospitalisation rates.

The third wave of infections in England due to Omicron was initially driven by BA.1, but by 10 January 2022, 53 sequences of the Omicron sublineage BA.2 had been identified in the UK^29^. BA.2 is widespread globally and has been phylogenetically divided into five groups^30^. A risk assessment for BA.2 published by the UK Health Security Agency (UKHSA) on 23 February 2022 reported, with high confidence, that BA.2 had a growth rate advantage compared to BA.1^31^. In round 17 of the REACT-1 study (between 5 and 20 January 2022) we reported a growth rate advantage for BA.2 compared to BA.1 and BA.1.1 (combined) with an additive R advantage of 0.46 (95% CI: 0.10–0.92), despite the small number (19) of BA.2 samples in the REACT-1 data at that time^14^. Our round 18 data based on sequencing of positive swabs and now with over 300 BA.2 cases strongly support this observation of a growth rate advantage for BA.2 compared with other Omicron sublineages. We estimate that by 21 February 2022, nearly 50% of all Omicron infections were BA.2. We also detected one instance of AY.39 Delta sublineage which may be a recent importation.

UKHSA has reported, with moderate confidence, that the transmissibility of BA.2 rather than its immune evasion were driving its growth rate advantage. This is supported by subsequent antigenic characterisation indicating that the BA.1 and BA.2 sublineages are antigenically equidistant from wildtype SARS-CoV-2^32^. In our own data, the lower Ct values (higher viral load) for BA.2 compared with BA.1 and BA.1.1 could indicate that BA.2 infections are more recent and/or that the higher viral loads are leading to greater transmissibility^33,34^. Furthermore, UKHSA reported, with moderate confidence, that the severity of BA.2 was similar to that of BA.1^31^, which is associated with lower rates of hospitalisation and deaths during the Omicron wave relative to those during the first wave (wildtype) and second wave (Alpha) in England^14^.

Our data indicate a geographical divide in BA.2 Omicron infections, with more infections due to BA.2 in the south compared to the north of the country. We found the highest proportion of BA.2 (among Omicron infections) in London where the initial Omicron epidemic in England took off in December 2021^4^. The highest prevalence of infection at local area level was also found in London. The reasons for this are unclear but may reflect higher levels of social interactions in the capital following announcements to end all domestic legal restrictions related to COVID-19, including a return to the workplace. Our observation of increased risk of infection among healthcare and care home workers reflects experience “on the ground” of high levels of absenteeism for COVID-19 among healthcare staff during the peak of the Omicron epidemic^35^.

Our study has limitations. We have observed a slow decline in response rates since the beginning of the study, from approximately 30% returning swabs with valid RT-PCR test results in round 1 (1 May to 1 June 2020) to 12.2% in round 17 (5 to 20 January 2022). To improve response rates and make the study more representative we included a small monetary incentive in round 18 among those aged 13 to 44 years, following a successful pilot. This had the result of more than doubling response rates at these ages, giving an overall response rate of 15.0%. We used rim weighting to correct the sample to the characteristics of the base population^16^. Unlike in previous rounds, the rim weighting correction led to only small changes in round 18 between the unweighted and weighted prevalence, reflecting the improved representativeness of the obtained sample.

In conclusion, we report continued high prevalence of SARS-CoV-2 infections during February 2022 as the Omicron wave in England has persisted. The ongoing replacement by BA.2 of other Omicron sublineages demonstrates a transmission advantage for BA.2 which may be contributing to the high rates of infection, alongside the opening up of society as all domestic legal restrictions related to COVID-19 in England were lifted. Of some concern is the recent uptick in hospitalisations in England which may reflect the level or increasing rates of infection in older people in the most recent data. Continued surveillance of infection and hospitalisation rates is required to evaluate whether these trends continue in the future.

## Data Availability

Access to REACT-1 individual-level data is restricted to protect participants anonymity.
Summary statistics, descriptive tables, and code from the current REACT-1 study are available at https://github.com/mrc-ide/reactidd (doi 10.5281/zenodo.6242826). REACT-1 study materials are available for each round at
https://www.imperial.ac.uk/medicine/research-and-impact/groups/react-study/react-1-study-materials/
Sequence read data are available without restriction from the European Nucleotide Archive at https://www.ebi.ac.uk/ena/browser/view/PRJEB37886, and consensus genome sequences are available from the Global initiative on sharing all influenza data (GISAID).

## Contributors

PE, CAD and MC-H are corresponding authors. PE, MC-H and CAD conceived the study and the analytical plan. MC-H, DT, OE, BB, HWang, JJ, DH, and CEW performed the statistical analyses. DT, HWang, OE, DH, BB, and MW, DH curated the data. JE, CA, PJD, DA, WB, GT, GC, HWard, AD provided study oversight and results interpretation. AJP and AJT generated the sequencing data. AD and PE obtained funding. All authors revised the manuscript for important intellectual content and approved the submission of the manuscript. PE, MC-H, CAD had full access to the data and take responsibility for the integrity of the data and the accuracy of the data analysis and for the decision to submit for publication.

## Acknowledgements

MC-H and BB acknowledge support from Cancer Research UK, Population Research Committee Project grant ‘Mechanomics’ (grant No 22184 to MC-H). MC-H acknowledges support from the H2020-EXPANSE (Horizon 2020 grant No 874627) and H2020-LongITools (Horizon 2020 grant No 874739). AJP acknowledges the support of the Biotechnology and Biological Sciences Research Council (BB/R012504/1). HWard acknowledges support from a National Institute for Health Research (NIHR) Senior Investigator Award, the Wellcome Trust (205456/Z/16/Z), and the NIHR Applied Research Collaboration (ARC) North West London. JE is an NIHR academic clinical fellow in infectious diseases. GC is supported by an NIHR Professorship. CAD acknowledges support from the MRC Centre for Global Infectious Disease Analysis, the NIHR Health Protection Research Unit in Emerging and Zoonotic Infections and the NIHR-funded Vaccine Efficacy Evaluation for Priority Emerging Diseases (PR-OD-1017-20007). PE is Director of the Medical Research Council (MRC) Centre for Environment and Health (MR/L01341X/1, MR/S019669/1). PE acknowledges support from Health Data Research UK (HDR UK); the NIHR Imperial Biomedical Research Centre; NIHR Health Protection Research Units in Chemical and Radiation Threats and Hazards, and Environmental Exposures and Health; the British Heart Foundation Centre for Research Excellence at Imperial College London (RE/18/4/34215); and the UK Dementia Research Institute at Imperial College London (MC_PC_17114). We thank key collaborators on this work – Ipsos MORI: Kelly Beaver, Sam Clemens, Gary Welch, Nicholas Gilby, Kelly Ward, Galini Pantelidou and Kevin Pickering; Institute of Global Health Innovation at Imperial College London: Gianluca Fontana, Justine Alford; School of Public Health, Imperial College London: Eric Johnson, Rob Elliott, Graham Blakoe; Quadram Institute, Norwich, UK: Nabil-Fareed Alikhan; North West London Pathology and Public Health England (now UKHSA) for help in calibration of the laboratory analyses; Patient Experience Research Centre at Imperial College London and the REACT Public Advisory Panel; NHS Digital for access to the NHS register; the Department of Health and Social Care for logistic support.

## Data sharing

Access to REACT-1 individual-level data is restricted to protect participants’ anonymity. Summary statistics, descriptive tables, and code from the current REACT-1 study are available at https://github.com/mrc-ide/reactidd (doi 10.5281/zenodo.6242826). REACT-1 study materials are available for each round at https://www.imperial.ac.uk/medicine/research-and-impact/groups/react-study/react-1-study-materials/

Sequence read data are available without restriction from the European Nucleotide Archive at https://www.ebi.ac.uk/ena/browser/view/PRJEB37886, and consensus genome sequences are available from the Global initiative on sharing all influenza data (GISAID).

## Supplementary material

**Table S1.** Unweighted and weighted prevalence of swab-positivity from REACT-1 across rounds 1 to 18.

| Round | Tested swabs | Positive swabs | Unweighted prevalence (95% CI) | Weighted prevalence (95% CI) | First sample | Last sample |
| --- | --- | --- | --- | --- | --- | --- |
| 1 | 120,620 | 159 | 0.13% (0.11%, 0.15%) | 0.16% (0.13%, 0.19%) | 01/05/20 | 01/06/20 |
| 2 | 159,199 | 123 | 0.08% (0.07%, 0.09%) | 0.09% (0.07%, 0.11%) | 19/06/20 | 07/07/20 |
| 3 | 162,821 | 54 | 0.03% (0.03%, 0.04%) | 0.04% (0.03%, 0.05%) | 24/07/20 | 11/08/20 |
| 4 | 154,325 | 137 | 0.09% (0.08%, 0.11%) | 0.13% (0.01%, 0.15%) | 20/08/20 | 08/09/20 |
| 5 | 174,949 | 824 | 0.47% (0.44%, 0.50%) | 0.60% (0.55%, 0.71%) | 18/09/20 | 05/10/20 |
| 6 | 160,175 | 1,732 | 1.08% (1.03%, 1.13%) | 1.30% (1.21%, 1.39%) | 16/10/20 | 02/11/20 |
| 7 | 168,181 | 1,299 | 0.77% (0.73%, 0.82%) | 0.94% (0.87%, 1.01%) | 13/11/20 | 03/12/20 |
| 8 | 167,642 | 2,282 | 1.36% (1.31%, 1.42%) | 1.57% (1.49%, 1.66%) | 06/01/21 | 22/01/21 |
| 9 | 165,456 | 689 | 0.42% (0.39%, 0.45%) | 0.49% (0.44%, 0.55%) | 04/02/21 | 23/02/21 |
| 10 | 140,844 | 227 | 0.16% (0.14%, 0.18%) | 0.20% (0.17%, 0.23%) | 11/03/21 | 30/03/21 |
| 11 | 127,408 | 115 | 0.09% (0.07%, 0.11%) | 0.10% (0.08%, 0.13%) | 15/04/21 | 03/05/21 |
| 12* | 108,911 | 135 | 0.12% (0.10%, 0.15%) | 0.15% (0.12%, 0.18%) | 20/05/21 | 07/06/21 |
| 13 | 98,233 | 527 | 0.54% (0.49%, 0.58%) | 0.63% (0.57%, 0.69%) | 24/06/21 | 12/07/21 |
| 14** | 100,527 | 764 | 0.76% (0.71%, 0.82%) | 0.83% (0.76%, 0.89%) | 09/09/21 | 27/09/21 |
| 15*** | 100,112 | 1,399 | 1.40% (1.33%, 1.47%) | 1.57% (1.48%, 1.66%) | 19/10/21 | 05/11/21 |
| 16**** | 97,089 | 1,192 | 1.23% (1.16%, 1.30%) | 1.41% (1.33%, 1.51%) | 23/11/21 | 14/12/21 |
| 17† | 102,174 | 4,073 | 3.99% (3.87%, 4.11%) | 4.41% (4.25%, 4.56%) | 05/01/22 | 20/01/22 |
| 18‡ | 94,950 | 2,731 | 2.88% (2.77%, 2.98%) | 2.88% (2.76%, 3.00%) | 08/02/22 | 01/03/22 |

\* Sampling strategy changed for round 12 and subsequent rounds. Therefore unweighted prevalence is not directly comparable with previous rounds
\*\* Including N=509 samples from 28-30 September 2021. Sample handling changed in round 14. Therefore prevalence is not directly comparable with previous rounds
\*\*\* Including N=93 samples (all negatives) from 6-8 November 2021, and N=86 samples with no collection/arrival dates
\*\*\*\* Including N=661 samples (including 12 positives ) from 15-17 December 2021. Swab positivity was assessed using a multiplex assay from round 16 onwards. Test diagnostic characteristics may differ slightly from previous rounds
† Including N=862 (including 36 positives) from 21-24 January 2022
‡ Including N=685 (including 18 positives) from 2-4 March 2022

**Table S2.**
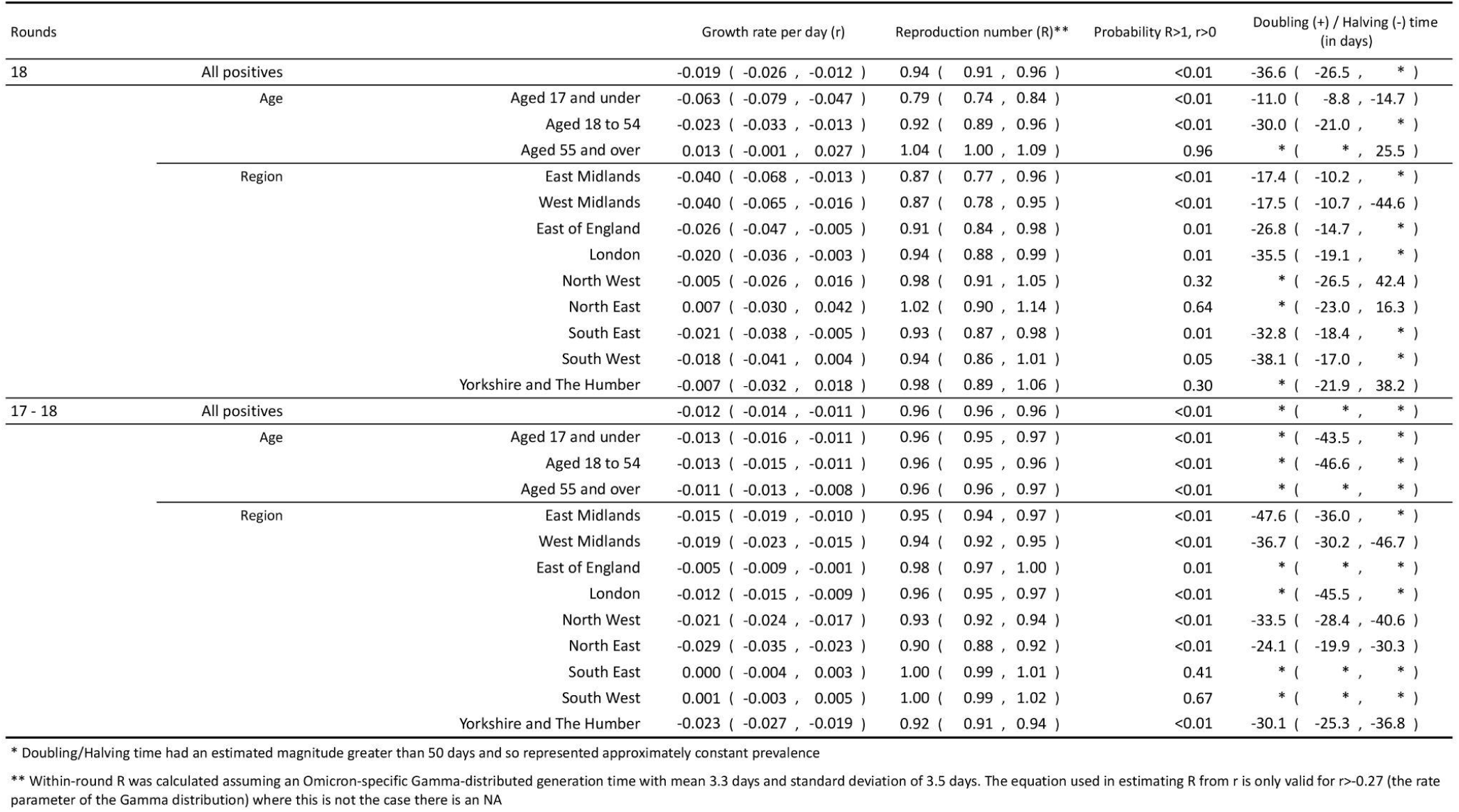
Table of growth rates, reproduction numbers and doubling/halving times from exponential model fits on data from round 17 (5 to 20 January 2022) and round 18 (8 February to 1 March 2022)

**Table S3A.**
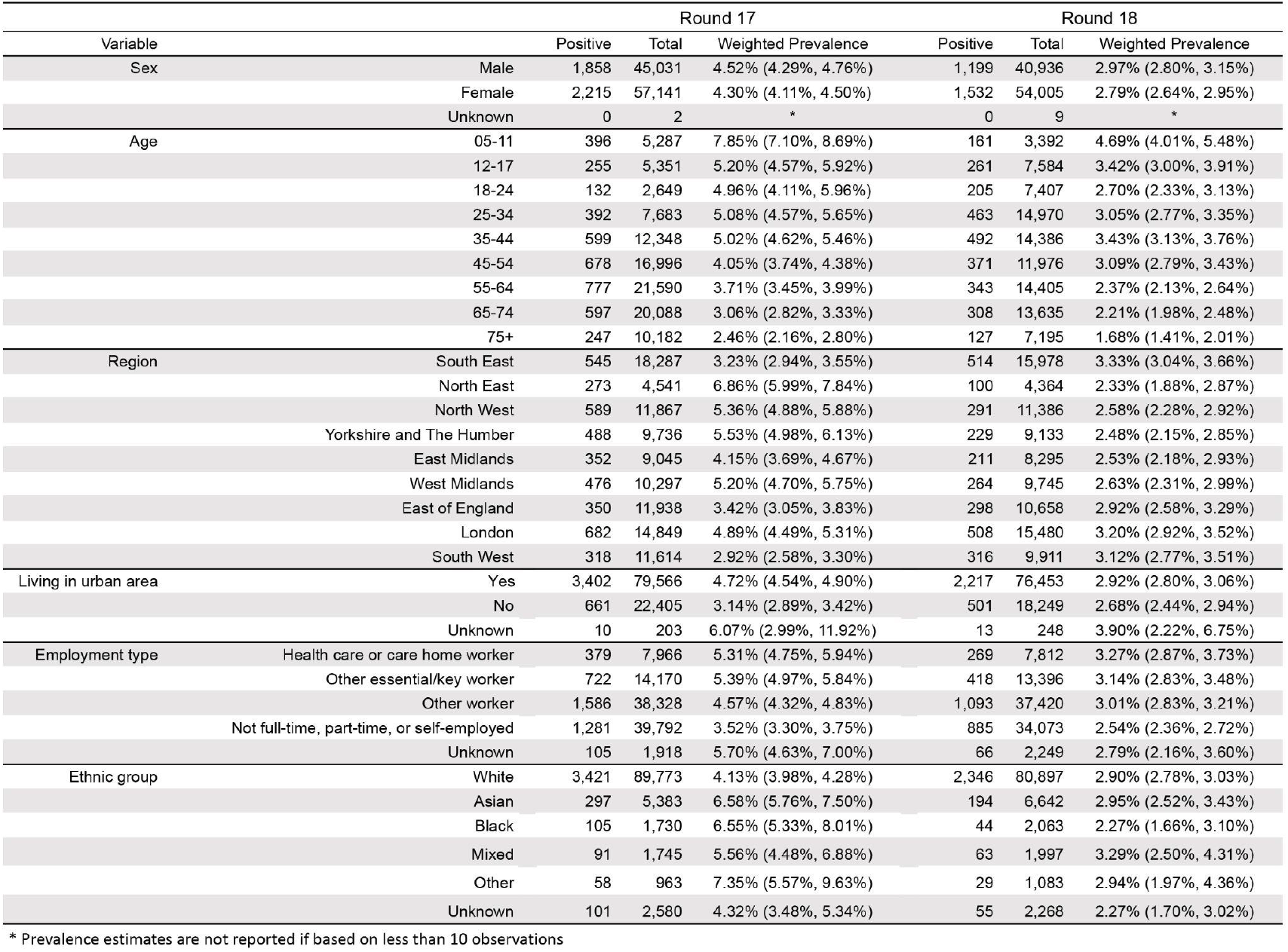
Weighted prevalence of SARS-CoV-2 swab-positivity in round 17 and round 18 by sex, age, region, urban status, employment type, and ethnic group.

**Table S3B.** Weighted prevalence of SARS-CoV-2 swab-positivity in round 17 and round 18 by household size, number of children in the household, contact with a COVID-19 case, protective behaviours, symptom status and neighbourhood deprivation.

|  |  | Round 17 |  |  | Round 18 |  |  |
| --- | --- | --- | --- | --- | --- | --- | --- |
| Variable |  | Positive | Total | Weighted Prevalence | Positive | Total | Weighted Prevalence |
| Household size | 1 | 536 | 17,601 | 3.15% (2.86%, 3.48%) | 288 | 14,239 | 1.87% (1.65%, 2.11%) |
|  | 2 | 1,304 | 40,719 | 3.38% (3.19%, 3.59%) | 845 | 33,450 | 2.50% (2.33%, 2.68%) |
|  | 3 | 786 | 17,384 | 4.95% (4.58%, 5.36%) | 543 | 18,194 | 3.06% (2.79%, 3.35%) |
|  | 4 | 941 | 18,422 | 5.41% (5.04%, 5.80%) | 716 | 19,042 | 3.88% (3.58%, 4.20%) |
|  | 5 | 337 | 5,719 | 6.23% (5.52%, 7.01%) | 219 | 6,570 | 3.33% (2.87%, 3.85%) |
|  | 6+ | 169 | 2,329 | 7.72% (6.47%, 9.18%) | 120 | 3,455 | 3.36% (2.76%, 4.09%) |
| Number of children in the household | 0 | 2,306 | 68,713 | 3.57% (3.41%, 3.74%) | 1,407 | 58,734 | 2.31% (2.19%, 2.44%) |
|  | 1+ | 1,535 | 28,206 | 5.92% (5.60%, 6.25%) | 1,072 | 28,474 | 3.88% (3.63%, 4.14%) |
|  | Unknown | 232 | 5,255 | 4.75% (4.14%, 5.45%) | 252 | 7,742 | 3.16% (2.78%, 3.60%) |
| COVID case contact | No | 1,605 | 75,859 | 2.40% (2.27%, 2.54%) | 1,187 | 69,708 | 1.73% (1.62%, 1.84%) |
|  | Yes, contact with a confirmed/tested COVID-19 case | 1,781 | 13,692 | 12.83% (12.20%, 13.49%) | 1,056 | 10,312 | 10.35% (9.72%, 11.02%) |
|  | Yes, contact with a suspected COVID-19 case | 230 | 2,562 | 9.38% (8.14%, 10.79%) | 142 | 2,098 | 6.86% (5.77%, 8.15%) |
|  | Unknown | 457 | 10,061 | 5.30% (4.78%, 5.86%) | 346 | 12,832 | 2.55% (2.27%, 2.85%) |
| Shielding | Yes | 2,809 | 26,223 | 3.44% (3.18%, 3.73%) | 426 | 17,277 | 2.39% (2.16%, 2.65%) |
|  | No | 801 | 65,800 | 4.60% (4.41%, 4.80%) | 1,953 | 64,720 | 3.08% (2.93%, 3.23%) |
|  | Unknown | 463 | 10,151 | 5.31% (4.80%, 5.88%) | 352 | 12,953 | 2.58% (2.31%, 2.89%) |
| Frequency wearing mask indoors | Always | 1,774 | 49,795 | 3.84% (3.64%, 4.05%) | 717 | 24,303 | 2.85% (2.64%, 3.08%) |
|  | Sometimes | 1,295 | 31,768 | 4.33% (4.07%, 4.61%) | 1,111 | 37,147 | 3.01% (2.83%, 3.20%) |
|  | Hardly ever | 131 | 2,695 | 5.22% (4.32%, 6.30%) | 109 | 3,901 | 2.89% (2.37%, 3.52%) |
|  | Never | 68 | 1,640 | 4.60% (3.43%, 6.15%) | 36 | 1,378 | 2.40% (1.70%, 3.40%) |
|  | Unknown | 805 | 16,276 | 5.77% (5.35%, 6.22%) | 758 | 28,221 | 2.77% (2.56%, 3.00%) |
| Symptom status | Classic COVID symptoms* | 1,673 | 10,481 | 15.85% (15.06%, 16.68%) | 1,175 | 7,845 | 15.06% (14.21%, 15.95%) |
|  | Other symptoms | 930 | 16,525 | 5.81% (5.40%, 6.24%) | 530 | 13,651 | 3.85% (3.52%, 4.22%) |
|  | No symptoms | 1,019 | 65,149 | 1.87% (1.74%, 2.01%) | 682 | 60,676 | 1.17% (1.08%, 1.27%) |
|  | Unknown | 451 | 10,019 | 5.26% (4.74%, 5.82%) | 344 | 12,778 | 2.53% (2.26%, 2.84%) |
| Deprivation | 1 Most deprived | 639 | 11,245 | 6.08% (5.58%, 6.61%) | 366 | 13,161 | 2.69% (2.41%, 3.00%) |
|  | 2 | 763 | 17,276 | 4.62% (4.27%, 4.99%) | 458 | 17,218 | 2.68% (2.42%, 2.95%) |
|  | 3 | 831 | 21,472 | 3.96% (3.68%, 4.26%) | 597 | 19,829 | 2.98% (2.73%, 3.24%) |
|  | 4 | 895 | 24,536 | 3.93% (3.66%, 4.22%) | 644 | 21,549 | 3.13% (2.88%, 3.40%) |
|  | 5 Least deprived | 945 | 27,645 | 3.69% (3.44%, 3.96%) | 666 | 23,193 | 2.90% (2.67%, 3.14%) |

\* Classic COVID symptoms: loss or change of sense of smell or taste, fever, new persistent cough

**Table S4.**
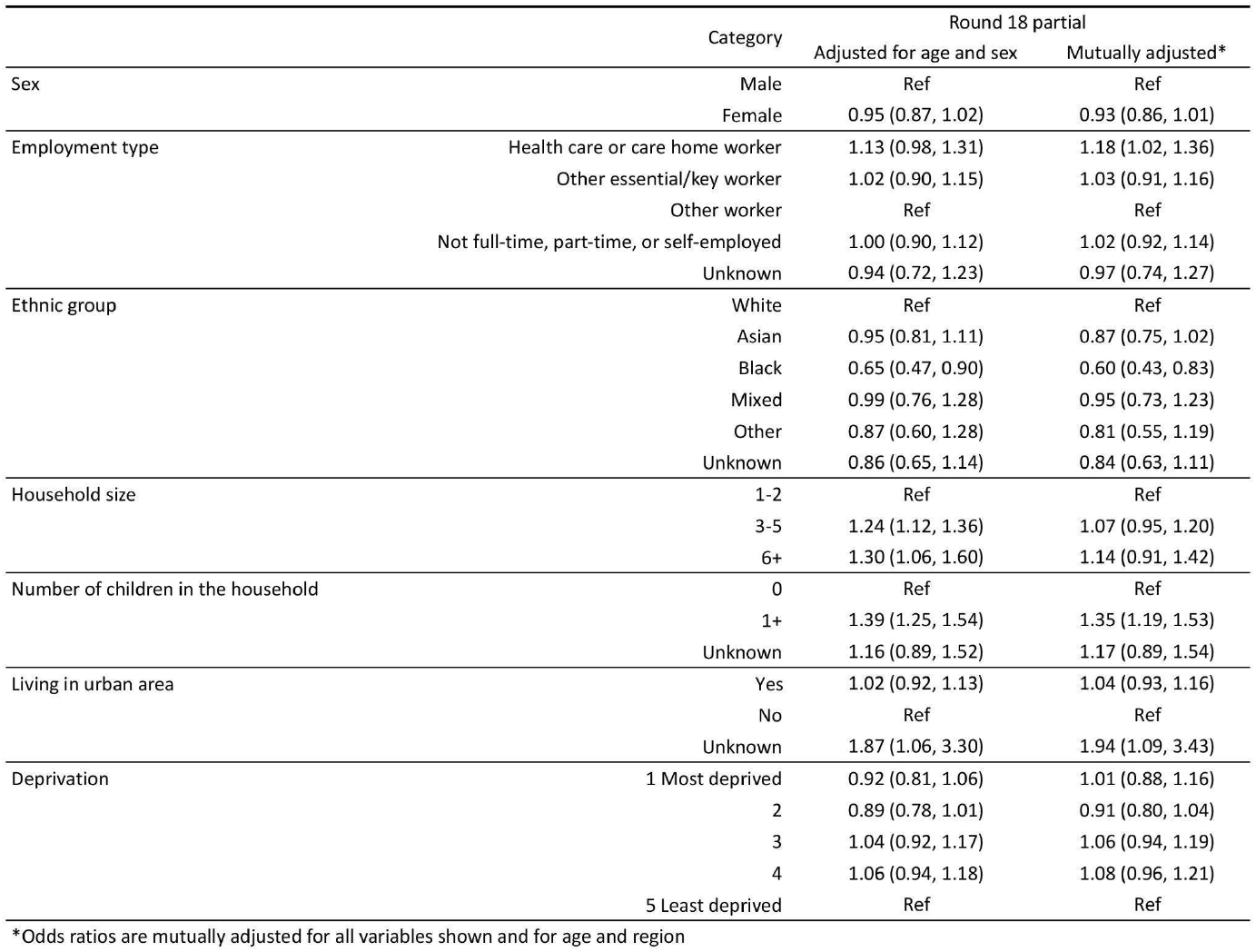
Multivariable logistic regression for round 18. Results are presented as Odds Ratios (95% confidence interval) adjusted for age and sex and additionally for region and all other variables (mutually adjusted OR).

|  | Category | Round 18 partial |  |
| --- | --- | --- | --- |
|  |  | Adjusted for age and sex | Mutually adjusted* |
| Sex | Male | Ref | Ref |
|  | Female | 0.95 (0.87, 1.02) | 0.93 (0.86, 1.01) |
| Employment type | Health care or care home worker | 1.13 (0.98, 1.31) | 1.18 (1.02, 1.36) |
|  | Other essential/key worker | 1.02 (0.90, 1.15) | 1.03 (0.91, 1.16) |
|  | Other worker | Ref | Ref |
|  | Not full-time, part-time, or self-employed | 1.00 (0.90, 1.12) | 1.02 (0.92, 1.14) |
|  | Unknown | 0.94 (0.72, 1.23) | 0.97 (0.74, 1.27) |
| Ethnic group | White | Ref | Ref |
|  | Asian | 0.95 (0.81, 1.11) | 0.87 (0.75, 1.02) |
|  | Black | 0.65 (0.47, 0.90) | 0.60 (0.43, 0.83) |
|  | Mixed | 0.99 (0.76, 1.28) | 0.95 (0.73, 1.23) |
|  | Other | 0.87 (0.60, 1.28) | 0.81 (0.55, 1.19) |
|  | Unknown | 0.86 (0.65, 1.14) | 0.84 (0.63, 1.11) |
| Household size | 1-2 | Ref | Ref |
|  | 3-5 | 1.24 (1.12, 1.36) | 1.07 (0.95, 1.20) |
|  | 6+ | 1.30 (1.06, 1.60) | 1.14 (0.91, 1.42) |
| Number of children in the household | 0 | Ref | Ref |
|  | 1+ | 1.39 (1.25, 1.54) | 1.35 (1.19, 1.53) |
|  | Unknown | 1.16 (0.89, 1.52) | 1.17 (0.89, 1.54) |
| Living in urban area | Yes | 1.02 (0.92, 1.13) | 1.04 (0.93, 1.16) |
|  | No | Ref | Ref |
|  | Unknown | 1.87 (1.06, 3.30) | 1.94 (1.09, 3.43) |
| Deprivation | 1 Most deprived | 0.92 (0.81, 1.06) | 1.01 (0.88, 1.16) |
|  | 2 | 0.89 (0.78, 1.01) | 0.91 (0.80, 1.04) |
|  | 3 | 1.04 (0.92, 1.17) | 1.06 (0.94, 1.19) |
|  | 4 | 1.06 (0.94, 1.18) | 1.08 (0.96, 1.21) |
|  | 5 Least deprived | Ref | Ref |

\*Odds ratios are mutually adjusted for all variables shown and for age and region

